# Proteomic analysis of extracellular vesicles in cerebrospinal fluid of patients with Alzheimer’s disease

**DOI:** 10.1101/2020.02.22.20026609

**Authors:** Davide Chiasserini, Irene Bijnsdorp, Giovanni Bellomo, Pier Luigi Orvietani, Sander R. Piersma, Thang V. Pham, Lucilla Parnetti, Connie R. Jiménez

## Abstract

Cerebrospinal fluid (CSF) contains different types of extracellular vesicles (EVs) with undisclosed biomarker potential for neurodegenerative diseases. The aims of the present study were: i) to compare the proteome EVs isolated using different ultracentrifugation speed ii) to preliminary explore the EVs proteome in a common neurodegenerative disorder, Alzheimer’s disease (AD) compared to neurological controls. CSF samples from control subjects and AD patients were pooled separately (15 mL) and subjected to ultracentrifugation (UC) at different speeds (20,000g and 100,000g) to isolate separate EV fractions (P20 and P100). The proteome was analysed using high-resolution mass spectrometry (LC-MS/MS) and comparisons were made using bioinformatic analysis. EVs isolated at 100,000g (P100) had a proteome consistent with vesicles secreted via an ESCRT-dependent mechanism, being highly enriched in alix (PDCD6IP), syntenin-1 (SDCBP) and TSG101. EVs isolated at 20,000g were substantially different, showing enrichment in cytoskeletal and cell adhesion molecules. The pools from patients diagnosed with AD showed a distinct protein profile of CSF EVs, with increased levels of ADAM10, SPON1, CH3IL1 and MDK in the P100 fraction. CSF EV offer a new potential biosource of protein markers for AD detection and a complementary framework to the analysis of whole biological fluids for biomarker discovery.

## Introduction

Extracellular vesicles (EVs) are membrane-enclosed vesicles secreted by a variety of cells and released in numerous biological fluids ^1^. Pleiotropic functions have been described for EVs, but a general consensus exists on their significant roles in signaling and information exchange between cells ^2,3^. Since intercellular communication is the defining characteristic of our central nervous system (CNS), several studies have investigated the role of EVs in the CNS, with particular consideration towards the transport of EV cargos, including proteins, lipids and nucleic acids and the effect on target cells ^4,5^. In addition, EVs in brain and cerebrospinal fluid (CSF) are attracting considerable attention for their possible role in the so-called prion hypothesis, the pathological spreading of protein aggregates across CNS structures, a characteristic feature of several neurodegenerative disorders like Alzheimer’s disease (AD) ^6,7^.

CSF represents a privileged sample to explore the function of EVs in CNS, since its composition may effectively mirror molecular changes related to neurodegeneration processes ^8,9^. EVs in CSF have been isolated with a variety of techniques to be characterized morphologically and biochemically ^10–12^. Among these techniques, differential ultracentrifugation is still considered one of the most reliable method for isolation of EV ^13^. Using differential centrifugation several subtypes of EVs have been described in both cell media and biological fluids ^14^. It has been shown that using lower centrifugation speed (10,000 or 20,000 x g) usually larger vesicle tend to be isolated, while at higher speed (100,000 x g), nanosized vesicle (< 150 nm) can be readily separated ^14^. These different types of vesicles may not only carry different protein cargos, but also have different physiological roles ^14^. To understand the precise molecular composition of these different type of vesicles may further support the studies on the function and therapeutic applications of EVs, currently limited by several factors including reliability of isolation techniques, low yield and contamination (especially in biological fluids) and lack of standardization ^13^. The proteome composition of the different types of EVs in CSF isolated with differential ultracentrifugation is not clearly defined yet and may support the existence of physiologically diverse CSF EVs, which can derive from different brain areas and be involved in a range of biological functions ^11^.

Recent efforts have been made to investigate the role of EVs in neurodegeneration and particularly in AD, either to clarify the involvement of EV in AD pathogenesis or to explore their value in diagnostics ^15^. It has been shown that EVs from AD brains contain amyloid beta (Aβ) oligomers that can propagate and induce cytotoxicity in vitro ^16^. Also the tau protein, a key player in AD pathogenesis, has been shown to spread via microglia-derived EVs, and stopping the production of these EVs contributed to the decrease of tau aggregation in vitro and in vivo ^17^. Additionally, several studies used immunochemical methods to analyse the biomarker potential of EVs in both plasma and CSF for AD diagnosis ^18–20^.

Here we used differential centrifugation, high-resolution liquid chromatography mass-spectrometry (LC-MS/MS) and bioinformatics analyses to characterise the proteomic landscape of CSF EVs isolated at 20,000 and 100,000 x g from pooled CSF samples of AD patients and control subjects. Our aims were: i) to delineate the core proteome of the different EVs in CSF and possibly define specific protein markers for each type of EV; ii) to analyse preliminarily the differential protein expression in AD EVs isolated from CSF pools. The final objective was to obtain a protein profile characteristic of AD EVs to be used in the development of new diagnostic biomarkers for AD, possibly complementing the core biochemical markers used in the clinic.

## Materials and methods

### CSF samples and EV isolation

CSF samples were obtained from patients that were retrospectively selected from the biobank of the Neurology Clinic of the University of Perugia. All the procedures involving human subjects were performed following Helsinki Declaration. All patients or their closest relatives gave informed consent for the inclusion in the study that was approved by the local Ethics Committee. All the patients included in the biobank underwent a baseline clinical examination by experienced neurologists, neuropsychological assessment using Mini Mental State Examination, blood chemistry, brain CT and/or MRI scan for excluding other causes of cognitive deficit. Lumbar puncture was performed between 8:00 and 10:00 a.m. CSF (10-12 mL), and was collected in sterile polypropylene tubes, centrifuged for 10 minutes at 2000 x g and divided in 0.5 mL aliquots which were immediately frozen at −80°C.

The AD pool was constructed using 30 CSF samples from patients diagnosed with probable AD according to Dubois et al [1]. The patients were selected so that the CSF profile of the classical AD biomarkers, amyloid β peptide 1-42 (Aβ42), total tau protein (t-tau) and phosphorylated tau 181 (p-tau) was supportive of AD (Aβ42 < 800 pg/mL, t-tau > 300 pg/mL, p-tau > 60 pg/mL) [2] (Figure S1). The control CSF pool was built from 30 subjects who underwent CSF tapping for diagnostic testing and, after clinical and biochemical evaluation, they were found free of any cognitive impairment, neurodegenerative or inflammatory disease (table S1). All control subjects had a negative AD biomarker profile. The CSF samples were thawed, pooled and immediately processed in order to limit the cycles of freeze and thawing to a single one.

EV isolation was performed as previously reported with minor modifications ^11^. The workflow of EV isolation is reported in figure 1a. The total pools (30 mL) of each group, CTRL and AD, were divided in two aliquots of 15 mL to have two technical replicates for the proteomic analysis. Spinning at low speed (1500-2000 x g) was omitted since all the samples were already spun at 2000 x g before freezing. Briefly, CSF samples were spun at 20,000 × g for 45 min in a ultracentrifuge using a 40Ti rotor (Beckman Coulter). The pellet was then washed with PBS and centrifuged again for 45 minutes at 20,000 x g to obtain the final sample (P20). The supernatant after 20,000 x g centrifugation was centrifuged at 100,000 × g for 2 hours. The soluble CSF (supernatant, SN) was removed and the pellet containing EVs were suspended in PBS, carefully washed and centrifuged again at 100,000 ×g for 2 hours to collect the final P100 pellet. All the centrifugation steps were performed at 4 °C. Pellets were dissolved in sample buffer (Life Technologies, Carlsbad, CA, cat No:NP0008) for proteomics experiments. SN fractions (∼5 mL) were concentrated to 100 µL using 3kDa filters (Millipore) and then dissolved in sample buffer.

**Figure 1.**
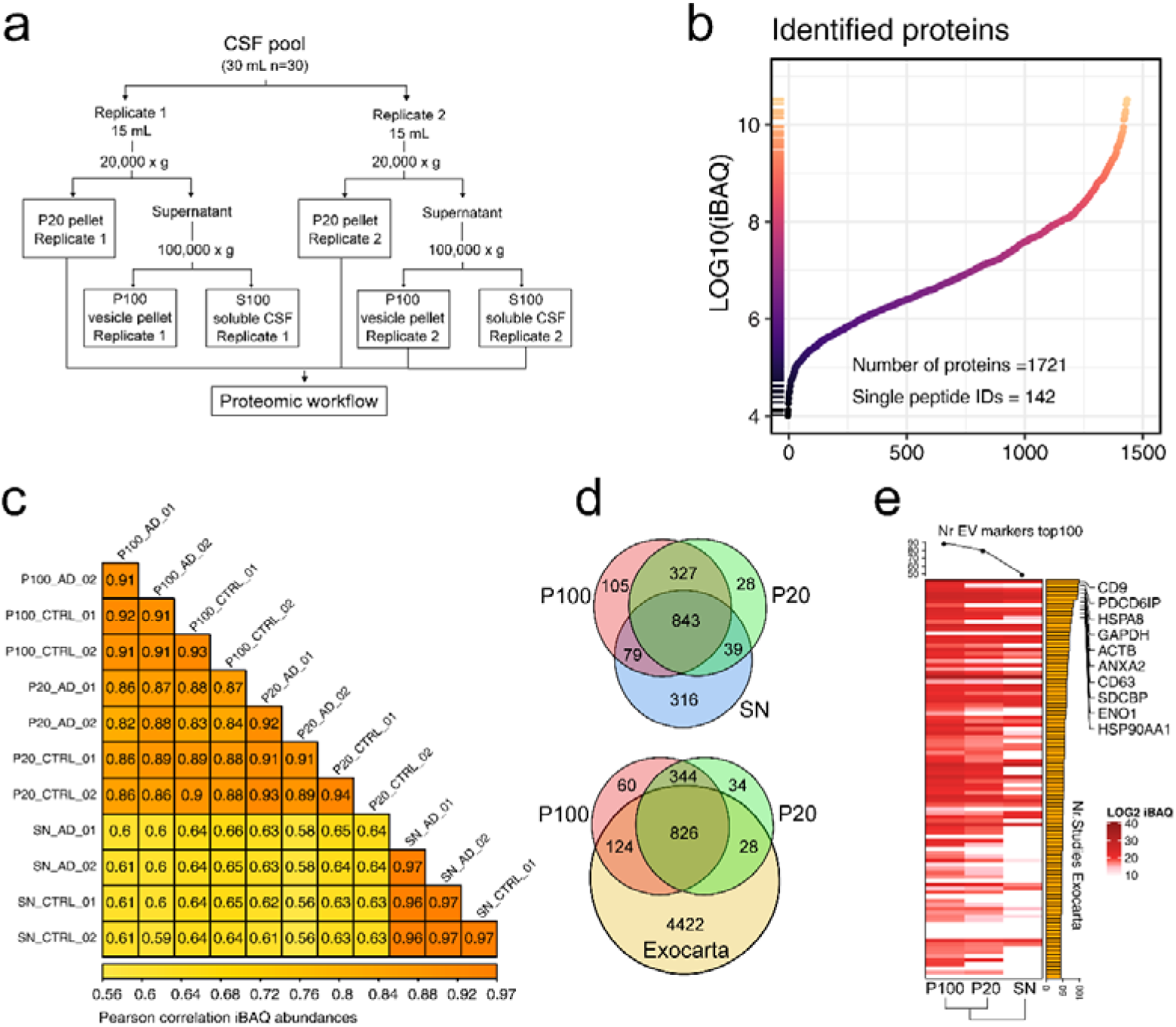
Characterization of CSF EV proteome. a) Scheme representing the experimental workflow for vesicle and supernatant (SN) isolation; b) scatter plot of iBAQ intensity values for all the identified proteins, 1760 proteins across 7 orders of magnitudes were identified in the whole dataset; c) Pearson correlation values of iBAQ abundances for each sample, technical replicates showed a very high correlation, in the range of 0.9 and 0.97, while the correlation of protein expression between SN and EV fractions was lower (∼0.6); d) Venn diagram of P100 vs P20 vs SN (upper graph) and P10 vs P20 vs whole Exocarta database. The proteins were considered as gene products and gene names were used for overlap. Only proteins identified in all samples were included in the analysis; e) heatmap of top-100 exosomes biomarkers in Exocarta (downloaded in December 2019) and their expression in the three experimental groups, according to iBAQ intensities. The bar graph on the right is relative to the number of studies in Exocarta identifying the protein, the dot plot on top of the heatmap is relative to the number of EV markers identified in each sample, with the P100 EVs showing the highest number of markers, followed by P20 and the supernatant (SN) fraction.

### SDS-PAGE and in-gel digestion

For proteomic analysis P100, P20 and SN fractions of the two pools were loaded on gradient gels from Invitrogen (NuPAGE 4–12% Bis-Tris gel, 1 mm × 10 wells). The gels were then stained with Coomassie brilliant blue G-250 (Pierce, Rockford, IL) and in-gel digested as previously described ^21^. Briefly, gels were washed and dehydrated once in 50 mM ammonium bicarbonate (ABC) and twice in 50 mM ABC/50 % acetonitrile (ACN). Cysteine bonds were reduced by incubation with 10 mM DTT/50 mM ABC at 56°C for 1 h, and alkylated with 50 mM iodoacetamide/50 mM ABC at room temperature (RT) for 45 minutes. After washing sequentially with ABC 50 mM and ABC/50% ACN, the whole gel was sliced in 5 bands for each lane. Gel parts were sliced up into approximately 1-mm cubes and collected in tubes, washed in ABC/ACN and dried in a vacuum centrifuge. Gel cubes were incubated overnight with 6.25 ng/mL trypsin and covered with ABC to allow digestion. Peptides were extracted once in 1% formic acid and twice in 5% formic acid/50% ACN. To remove gel particles and contaminants prior to LC-MS analysis, the volume of the peptide extract was reduced to 60 µL in a vacuum centrifuge and filtered using a 0.45 µm spin filter. Peptides were then resuspended in 20 ul of loading buffer (2 % ACN / 0.1 % formic acid).

### NanoLC-MS/MS analysis and database searching

Extracted peptides were separated on a 75 µm x 42 cm custom packed Reprosil C18 aqua column (1.9 µm, 120 Å) in a 90 min. gradient (2-32% Acetonitrile + 0.5% Acetic acid at 300 nl/min) using a U3000 RSLC high pressure nano-LC (Dionex). Eluting peptides were measured on-line by a Q Exactive mass spectrometer (ThermoFisher Scientific) operating in data-dependent acquisition mode. Peptides were ionized using a stainless-steel emitter at a potential of +2 kV (ThermoScientific). Intact peptide ions were detected at a resolution of 35,000 (at m/z 200) and fragment ions at a resolution of 17,500 (at m/z 200); the MS mass range was 350-1,500 Da. AGC Target settings for MS were 3E6 charges and for MS/MS 2E5 charges. Peptides were selected for Higher-energy C-trap dissociation fragmentation at an underfill ratio of 1% and a quadrupole isolation window of 1.5 Da, peptides were fragmented at a normalized collision energy of 30. Raw files from MS analysis were processed using the MaxQuant computational proteomics platform version 1.6.10.43 22. MS/MS spectra were searched against the Uniprot human database (release 2019_10, 20,394 sequences including isoforms) with initial precursor and fragment mass tolerance set to 7 and 20 p.p.m., respectively. Peptides with minimum of seven amino-acid length were considered with both the peptide and protein false discovery rate (FDR) set to 1%. Enzyme specificity was set to trypsin and up to two missed cleavage sites were allowed. Cysteine carbamidomethylation (Cys, +57.021464 Da) was searched as a fixed modification, whereas N-acetylation of proteins (N-terminal, +42.010565 Da) and oxidized methionine (Met, +15.994915 Da) were searched as variable modifications.

### Statistical and bioinformatic analyses

Statistical analysis and plotting were performed using the R language ^23^. Protein groups file from MaxQuant analysis was analysed using in-house scripts on label-free iBAQ intensities ^24^. This measure of protein abundance takes into account the number of theoretical peptides of each protein, giving an estimation of protein copy number ^24^, and allows a comparison of protein abundances within the specific sample. Limma R package ^25^ was used to find differentially expressed proteins (at least ± 1.5 fold change and p-value < 0.05). To find proteins enriched in the EV fractions we compared the three groups using a paired blocking design, while for the comparison AD vs CTRL, the only factor was the diagnostic group, with separate analyses for each type of sample. For quantification purposes, protein groups were filtered excluding all entries with an average of peptide-spectral matches (PSMs) across the samples less than 1, this initial filtering allowed the exclusion of very low abundance proteins before submitting the data to the limma analysis. Data were imputed using the MinProb method embedded in the imputeLCMD R package ^26^. Gene ontologies were analysed with the R package clusterProfiler ^27^ while gene sets were downloaded from the Molecular Signatures Database v7.0 ^28,29^. To obtain functional networks of the identified proteins we used the STRING database ^30^, using only experimental and knowledge-based physical interactions. The interaction network were exported and visualized in Cytoscape (v 2.8.3), using the combined score from STRING to describe the interaction strength ^31^. The final network was clustered using the MCL algorithm ^32^ embedded in clusterMaker Cytoscape plugin ^33^.

## Results

### Characterization of the EVs proteome

EVs isolated at different centrifugation speed were first characterized using SDS-PAGE. The protein pattern of EVs isolated at 100,000g was very similar to what obtained previously ^11^. The SDS-PAGE protein pattern was relatively similar between P100 and P20 (figure S2), while the SN fractions (30 µg) resembled the proteome of unfractionated CSF, with albumin as the most abundant protein band in the samples (figure S2).

Mass spectrometry analysis of gel-fractionated EV and SN fractions of the two CSF pools from AD and CTRL patients identified a total of 1721 protein groups (FDR < 1% both at peptide and protein level, figure 1B, table S2), corresponding to 1739 unique gene products. Proteins identified with single peptides were less than 1% (n=142). The overlap in protein identification among the replicates was high, ranging from a minimum of 76% in the P20 of the CTRL pool to the 86% of the SN fraction in both pools. This was confirmed also by correlation analysis (figure 1c), where the correlation of technical replicates ranged between 0.90 and 0.97.

About 50% of the proteins were shared between the three fractions (Figure 1 d). Interestingly 460 proteins were not identified in the soluble CSF (SN), but only in the P100 or P20 vesicle fractions. When the P100 and P20 fractions were compared with the proteins included in the online vesicle database Exocarta, we found 439 proteins never identified before in EVs, of these, 110 were specifically identified in both EV fractions and not the in the CSF SN (data not shown). The P100 EVs were the sample with the highest number of proteins included in the top 100 most commonly identified proteins in exosomes-like vesicles (Exocarta database, figure 1e). On average, the EV markers identified in the P100 fraction showed also the highest intensity as measured by iBAQ quantification when compared to the P20 and SN fractions (figure 1e), confirming the quantitative enrichment of EV proteins in the P100 and P20 fractions.

### Comparison of P100 and P20 EVs proteomes

Identification of EV specific proteins in biological fluids is hindered by the presence of high abundance proteins, which might be co-isolated ^34^. Here we used identification rate, differential expression analysis and network analysis to investigate the CSF EVs protein expression data and define a core proteome for both EV types. The limma statistical framework was applied to iBAQ protein abundances to calculate fold enrichment and statistical significance of the enrichment (table S3). Proteins with a fold change of at least 1.5 compared to the SN and identified in all four EV samples independently from the diagnostic group were considered identified in the specific type of vesicle.

Furthermore, the fold change of P100 vs P20 was used to discriminate proteins highly enriched for each vesicle type. Using this approach, 144 proteins were identified in the P100 fraction compared to the SN. Of these, 89 were specifically enriched also compared to the P20 fraction. Figure 2a shows a scatter plot of the normalized iBAQ intensities of the 144 identified P100 proteins. The highest expression was found for the sodium/iodide cotransporter (SLC5A5), Cysteine-rich and transmembrane domain-containing protein 1 (CYSTM1), CD9, lactadherin (MFGE8) and for the neuronal membrane glycoprotein M6-a (GPM6A). To further characterize the CSF EVs proteome we also applied network analysis to understand if functionally interacting proteins were identified in the enriched proteins lists. Figure 2b shows a network of the enriched P100 proteins (n=89) clustered according to interaction strength from STRING database and subjected to functional enrichment. This core proteome is characterized by the presence of EV markers (PDCD6IP, SDCBP, CD81, CD9) but shows also peculiarities possibly related to the type of biofluid/tissue with inclusion of a cluster of G proteins, transporters (components of the sodium/potassium ATPases) and ciliary/transport proteins (IFT74, IFT57, IFT172). The enrichment in EV markers in the P100 EVs confirms previous results ^11^ (figure 2b, the complete network is reported in figure S3). In the P20 fraction, 102 proteins were found enriched compared to the SN fraction while 51 were enriched also compared to the P100. The matrix-gla protein (MGP), pleiotrophin (PTN) and two S100 proteins (S100A8, S100A9) were highly expressed in the P20 fraction (figure 2c). The P20 specific proteins were dominated by a cluster of molecules involved in cell adhesion, and two small clusters involved in cell division and nucleosome assembly (figure 2d, figure S3).

**Figure 2.**
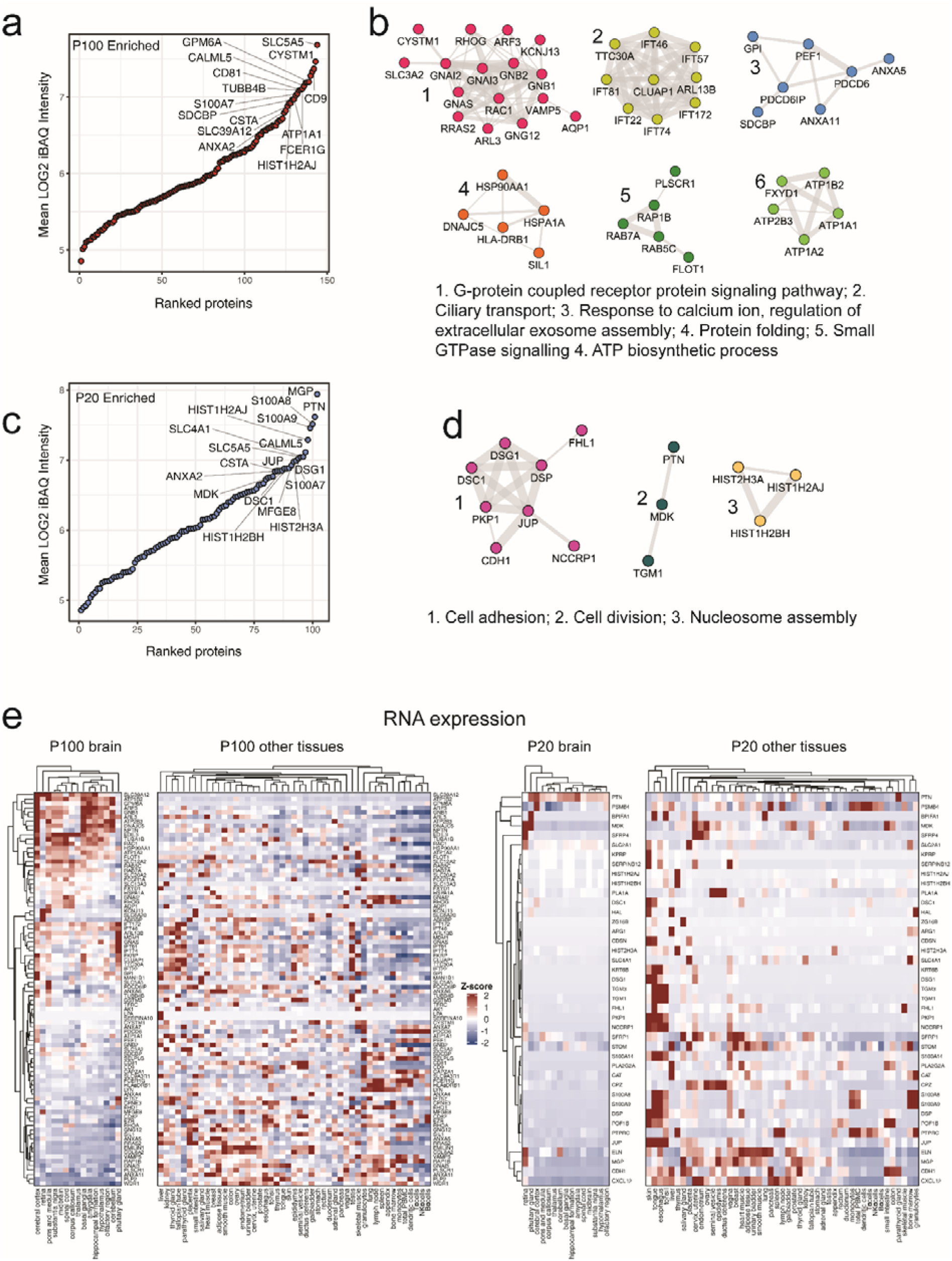
The proteome of CSF EVs isolated with differential centrifugation. a) Abundances (iBAQ intensities) of all the proteins identified in the P100 EV. Highest abundance were obtained for SLC5A5, CYSTM1, CD9, GPM6A; b) Top sub-networks for the P100 enriched proteins, the interaction network from the STRING database evidenced enrichment in G-proteins, ciliary proteins, exosome assembly, protein folding. A complete network is reported in figure S3. Circles are proteins indicated with gene name while edges are relative to the strength of the interaction according to the combined score of the STRING database; c) Protein abundances for the identified proteins in the P20 EVs, MGP, PTN and S100 proteins were the most abundant proteins according to iBAQ normalized intensities; d) Top sub-networks for the P20 enriched proteins. Main modules were related to cell adhesion, cell division and nucleosome assembly. Also for this panel the total network is reported in figure S3; e) Heatmap of P100 and P20 enriched proteins according to human protein atlas RNA expression data. Among the highly expressed proteins in brain structures, SLC39A12 and GPM6A enriches in P100 EVs, showed very low transcript expression in other tissues, in P20 EVs PTN showed high expression in CNS structures but also high expression in non-CNS tissues.

EVs within CSF contain a large number of brain-derived proteins, possibly secreted by different brain areas ^11^. However, a fraction of CSF-EVs might be derived from blood or even from other organs, due to extravasation of CSF circulation into plasma. The identification of specific markers of CSF EVs is therefore of fundamental importance to correctly characterize brain-derived EVs. Figure 2e shows the data clustering of proteins that are enriched in the P100 and P20 fractions against their RNA expression in several tissues, as reported in the Human Proteome Atlas ^35^. The RNA consensus data from three different sources (HPA, GTEx and FANTOM5) were used. After subdivision of the expression data in CNS-derived tissues and tissues or cells of other origin, we found that P100-enriched proteins showed a cluster of proteins highly enriched in CNS tissues compared to others. Among these, the neuronal membrane glycoprotein M6-a (GPM6A) was peculiarly enriched in the cerebral cortex and other brain structures while having very low expression values in non-CNS tissues. Similar results were obtained for the zinc transporter ZIP12 (SLC39A12). These two proteins may represent specific markers for exosomal (nano)vesicle in CSF. In the P20 fraction, only pleiotrophin (PTN) showed sustained RNA expression level in all the CNS-derived tissues, being expressed also in several non-CNS tissues. Interestingly, midkine (MDK), a protein with similar functions to PTN, was enriched in the P20 fraction and showed non-specific expression across human tissues.

### Differential protein expression in AD vs CTRL pools

Several studies investigated the biomarker potential of CSF EVs for neurological diseases, including AD ^36–38^. Here, we preliminary examined if the two types of vesicles isolated from pooled samples could carry AD protein biomarkers. To find differentially expressed proteins we used the limma statistical framework on P100 and P20 EV protein abundances and compared the AD vs CTRL pools. In the P100 AD vs P100 CTRL comparison we found 90 differentially expressed proteins (1.5 < fold change < −1.5), of which 71 up-regulated and 19 down-regulated in the AD pools. In the P20 fraction comparison, we found 122 differentially expressed proteins, with 103 being up-regulated and 19 down-regulated in AD vs CTRL pools (table S4). Interestingly, most of the differentially expressed proteins were specific of either one type of EV or the other, with an overlap of 18 proteins (figure 3a, b). Gene-set analysis using gene ontology categories for biological processes evidenced enrichment in different processes for differentially regulated proteins in P100 and P20. Up-regulated proteins in P100 showed enrichment in gene sets linked to lymphocyte migration, cell junctions and intraciliary protein transport, while down-regulated proteins were involved in defense response to exogenous organisms and morphology and cell polarity (figure S4). On the contrary, P20 up-regulated proteins were linked to lymphocyte activation or immune response processes regulating cell surface receptors, while down-regulated proteins showed only a single significant association with the gene set Golgi vesicle mediated transport (figure S4). To categorize further putative candidate biomarkers we investigated if differentially expressed proteins in CSF EVs were expressed in brain. We used two studies presenting proteomic data in brain areas important in AD pathogenesis, the hippocampus and the cortex (frontal gyrus). The study of Hondius and colleagues investigated protein expression in laser capture microdissected hippocampal tissue from patients at all pathologic stages of AD (Braak stages) ^39^, while the study of Bai and colleagues is a deep proteomic profiling of AD patients (frontal gyrus) using multi-omics techniques and data integration ^40^. A first overlap analysis evidenced that, globally, most of the differentially expressed proteins in CSF EVs are expressed either in the frontal gyrus (∼12,000 gene products) or hippocampal tissue (∼3000 gene products) (figure S5), with only 11 proteins not identified in brain for the P100 and 12 proteins for the P20. Secondly, we prioritized the putative candidates using a ranked list of AD–associated proteins assembled from different sources including the two brain datasets used for the overlap (see supplementary methods). Figure 3c shows the network of interactions of the differentially expressed proteins in EVs after ranking and filtering according to the number of evidences from the AD association list. We found that sixteen differentially expressed P100 proteins showed at least one other association with AD, while for the P20 proteins this number was equal to 22. Six proteins were significantly differentially expressed in AD pools in both type of EVs and were associated with AD.

**Figure 3.**
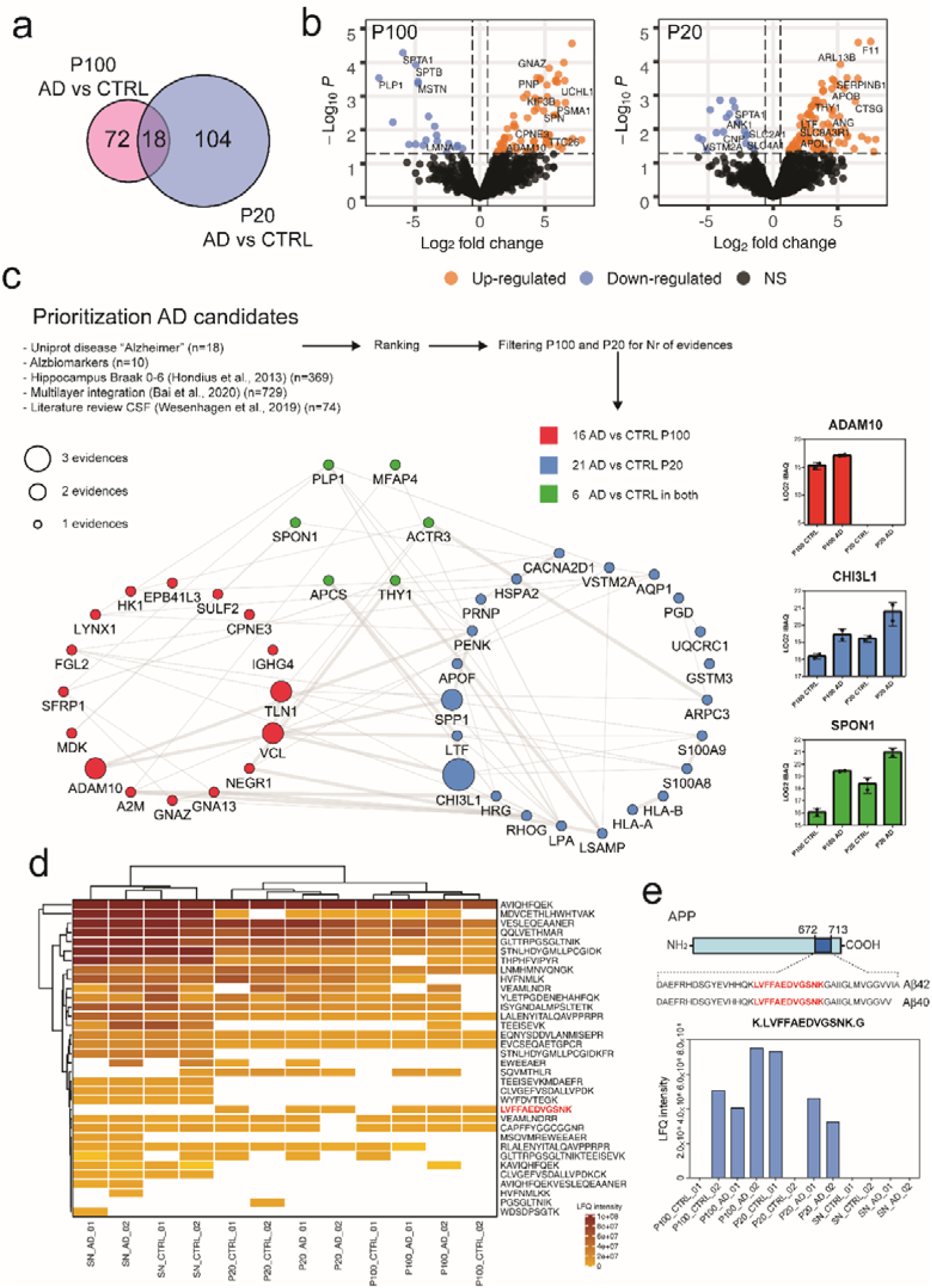
Differential protein expression in CSF EV between AD and control pools. **a)** Venn diagram of differentially expressed proteins in P100 and P20 EV fractions in CSF; b) Volcano plot of differentially expressed proteins for the AD vs CTRL comparison. In both vesicle types the majority of proteins was up-regulated in AD pools (fold change > 1.5 p-value <0.05); c) Prioritization of AD candidate biomarkers in CSF EVs visualized as interaction network. A list of AD-associated proteins was assembled (see supplementary methods) using experimental data, literature survey and knowledge-based evidence (UniProt database). Proteins showing significant changes in CSF EVs were ranked and filtered for the number of positive association with AD. Sixteen proteins from P100 EVs showed at least one positive association, while for the P20 EVs there were 21 proteins. Six proteins with positive association were significantly changed in both P100 and P20. ADAM10 and CHI3L1 showed the highest ranking among the differentially expressed proteins. On the right a barplot of three exemplary proteins for each of the three classes is included, values are reported as mean ± standard deviation; d) heatmap of label free quantification (LFQ) values of the amyloid precursor protein (APP) in CSF EVs and supernatants (SN). Values are normalized according to the MaxLFQ algorithm of the MaxQuant software; e) schematic of APP proteins and location of Aβ42 and Aβ40 in the APP sequence. Below the scheme, a barplot of the LFQ intensities of the Aβ peptide K.LVFFAEDVGSNK.G across the samples.

Among the proteins with higher number of knowledge-based and literature associations, ADAM10 was significantly overexpressed in the AD pools while it was not identified in the P20 EVs. Chitinase 3 like proteins 1 (CHI3L1 or YKL-40), a secreted inflammatory protein previously proposed as biomarker for AD ^41^, was significantly increased in the P20 EV and showed an increasing trend also in P100 EV (p = 0.069). Spondin-1 (SPON1) a glycoprotein involved in cell adhesion and with a genetic association with AD severity ^42^ was instead increased in AD pools both in P100 and P20 EVs (figure 3d). These results confirm that CSF EV contains AD-related proteins with biomarker value for AD detection.

Some studies investigated the presence of the core AD biomarkers (amyloid beta - Aβ - peptides and tau proteins) in EV from CSF or plasma from AD patients, mostly with immunological methods ^36^. To understand if AD biomarkers were present in CSF EVs we looked specifically for tau proteins and amyloid precursor protein (APP) peptides. While tau proteins were not identified in our dataset, neither in the supernatant nor in the EV fractions, several APP peptides were quantified with our proteomic approach, both in the SN and EV fraction. Figure 3d shows a heatmap of MaxQuant label free quantification (LFQ) for the APP peptides; in total 34 peptides were identified across the samples, with the majority identified in the SN fractions (also with higher intensity). All the APP peptides belonged to the extracellular domain of the protein (figure S6). While it is not possible to identify the Aβ isoforms having established diagnostic and pathological value like Aβ42 or Aβ40 ^43^ using our general proteomic approach (identification of tryptic peptides only), we identified the peptide LVFFAEDVGSNK situated around the middle of the Aβ sequence, and having tryptic termini. This peptide is included in both Aβ42 and Aβ40 and was identified and quantified in both P100 and P20 EV fractions, while it was not identified in the SN (figure 3e). While this peptide was found in both replicates of the AD P20 and P100 EVs fractions, it was found only in one of two replicates in control P20 and P100 fractions (figure 3e).

## Discussion

In this study, we investigated the proteomic landscape of EVs isolated from CSF using differential ultracentrifugation. The results show that using differential centrifugation speeds it is possible to isolate EVs from CSF with different protein cargo abundances. We identified specific marker of each type of vesicle and evidenced a differential protein profile between AD and CTRL EVs isolated from CSF.

We found that EVs isolated at different centrifugation speed carry, globally, a relatively similar proteome (80% protein ID overlap, figure 1d) but the protein abundances were strikingly different. The fraction obtained at 100,000 x g (P100) showed enrichment in a series of EV markers linked to the Endosomal Sorting Complex Required for Transport (ESCRT) machinery, including PDCD6IP, SDCBP and TSG101, but also endosomal markers as CD63, CD81, CD9, or other proteins like the disintegrin and metalloproteinase domain-containing protein 10 (ADAM10). Our results confirms the data from a previous a comprehensive study of proteins markers in EV isolated at different speed in cell supernatants, including the role of ADAM10 as marker of EV isolated at 100,000 x g ^14^. The ESCRT pathway, a conserved molecular machinery involved in the formation of multivesicular bodies (MVEs) ^44^, is comprised of about 30 proteins and has been connected to the biogenesis of EVs ^45^. Accordingly, we can propose that using high-speed centrifugation the resulting EV fraction maybe enriched in vesicles with ESCRT-dependent biogenesis.

The P100 EVs showed also an enrichment in other proteins classes, like G-proteins, component of the cilium and proteins involved in the ATP biosynthetic process (figure 2b). The enrichment in ciliary proteins is particularly interesting, since the main site for CSF production in the brain is the ciliate epithelium of the choroid plexus (CP) ^46,47^. Several components of the intraflagellar transport (IFT) complex, a protein machinery mediating the transport of proteins within the cilia and involved in ciliogenesis ^48^, were highly enriched in P100 EVs. Beside the IFT components, the enrichment of ciliary proteins in P100 EVs is confirmed also by the identification of other structural components of the cilia, like the small G proteins ADP-ribosylation factor like 3 (ARL3) and ADP ribosylation factor like GTPase 13B (ARL13B) ^49,50^. It has been shown that CP can produce EVs with a precise functional role in the regulation of communication between the peripheral systems and the brain, since the CP cells can sense peripheral inflammation and elicit secretion of EVs in CSF, to target different brain areas ^51^. The finding of ciliary proteins in our dataset may imply that a considerable part of EVs present in CSF is produced by the CP, and, bearing in mind the role of CP EV in communicating an inflammation state, EV in CSF may represent an interesting reservoir of molecules to detect CNS inflammation ^51^.

On the other hand, the P20 EVs showed enrichment in adhesion molecules like desmoglein 1 (DSG1), desmocollin (DSC1) and junction plakoglobin (JUP), belonging to the cadherins family ^52^. These molecules are present in the adherent junctions of the endothelial cell composing the blood-brain barrier ^53^. The occurrence of these adhesion molecules in the EV isolated at 20,000 x g might be thus exploited as a proxy to monitor BBB disruption ^54^.

The second module of enriched proteins in P20 EV included PTN and MDK, two growth factors involved in a variety of function in the CNS ^55^.

Our analysis of RNA expression data from the HPA showed that the majority of the proteins identified in EVs had a wide range of expression across human tissues (figure 2e). However, a cluster of proteins highly expressed in CNS structures and having negligible expression in other tissues clearly emerged from our data. Among these proteins, GPM6A, a membrane glycoprotein involved in neural differentiation ^56^ showed high expression in the cerebral cortex, basal ganglia and cerebellum. This molecule has been recently reported as differentially expressed in EV derived from AD brain tissues ^57^. In our data, GPM6A was not included in our candidate list, but showed an increasing trend in AD P100 EVs (table S4). Interestingly, we identified 6 peptides belonging to GPM6A, in both the extracellular and intracellular parts of the protein sequence (figure S6), supporting the hypothesis of the brain origin of this protein and its active inclusion in CSF EVs. The specific expression in brain tissue, together with the transmembrane properties of GPM6A make it an attractive target for immune-isolation of brain-derived EV in biological fluids, comparing favorably to other previously used protein markers as L1CAM ^19^.

Our preliminary analysis on differential protein expression in CSF EVs from AD patients and control subjects yielded a list of candidate molecules to be validated in following studies. ADAM10 was identified in P100 EVs and supernatants but not in the P20 EVs, and was significantly increased in EVs from AD patients. A recent study in human CSF reported ADAM10 levels as decreased AD patients compared to control subject ^58^. Interestingly, several forms of ADAM10 were found in CSF using immunoblotting, including unprocessed forms (range 55-80 kDa) and a soluble form possibly released from the membrane (∼50 kDa); the latter showing the reported decreased in AD CSF. In our dataset, we identified only peptides relatively close to the N-terminal and belonging to the soluble extracellular part of ADAM10. It would be interesting to speculate that differential distribution of ADAM10 between the soluble CSF and EVs may happen during the secretion process, an event that may explain the contrasting results obtained in this study when compared to the report by Sogorb-Esteve and colleagues ^58^.

In the P20 EVs, CHI3L1 (also called YKL40); an inflammatory glial protein, was differentially expressed in AD patients. This protein has been previously reported as CSF biomarker for AD by several authors ^59,60^, though with a diagnostic performance inferior to the core AD biomarkers ^41^. CHI3L1 is considered a marker of astrocytes activity ^61^, and its expression levels in CSF seem to be altered not only in AD but also in frontotemporal dementia with TDP43 mutations ^62^ and are associated with a greater atrophy rate in elderly subject without cognitive decline or dementia ^60^. Our data show a non-specific inclusion of CHI3L1 in both P100 and P20 CSF EVs and increased levels in AD patients; these results confirm the role of CHI3L1 as an AD biomarker, and the fact the CSF EV may mirror pathological events in AD brains.

The biomarker potential of CSF EVs is further showed by the finding of other two proteins with significant changes in AD EVs that have been linked to AD pathogenesis, spondin-1 (SPON1) and midkine (MDK). SPON1 is a cell adhesion protein possibly involved in axonal guidance and neuron polarization ^63^. It has been shown that SPON1 polymorphisms may affect brain structure and influence the risk for the onset of neurodegeneration processes in adult life ^42^. To our knowledge this report is the first one to find changes in CSF of AD patients for the SPON1 protein. MDK, on the other hand, has been shown to have a neurite growth promoting activity, and altered expression levels in AD brains have been reported before ^40^, as well as its association with plaque pathology ^64^.

The analysis of APP peptides in EV fractions showed the presence of a tryptic peptide belonging to the Aβ peptide (figure 3 d,e). This fragment has been found in both replicates of the P20 and P100 EVs fractions belonging to the AD CSF pools and only in one of two replicates in the control P20 and P100 EVs fractions. However, due the limited number of samples, it was not possible to validate the differntial expression of the Aβ peptide between AD and control pools. Nevertheless, this finding confirms that amyloidogenic peptides are present in EVs isolated from human CSF and may warrant further investigation with orthogonal techniques to understand their biomarker potential and their role in diseae spreading ^65^.

Our study has some limitations. Due to the relatively limited yield of EVs from CSF we used pooled samples to have enough proteins to characterize the protein expression of CSF EVs. This strategy may influence the significance analysis of our results, averaging the expression data from the patients included in the pools but also having positive effect as the normalization of biological variation ^66^. Accordingly, the pooling strategy has been succefully applied in other discovery proteomic studies of human brain where the paucity of material and biological variability are also relevant ^40,67^. The development of microfluid devices, single-molecule immunoassays and specific affinity reagents, together with the discovery of specific markers for different EV types will allow the isolation and sensitive detection of protein markers in clinically available volumes of biofluids from single patients ^68–70^.

In conclusion, EVs isolated from CSF using differential centrifugation have different protein markers that may be used to target specific vesicles or to isolate neuronal-derived vesicles from other biological fluids like plasma. The study of EVs in biological fluids may offer new exciting opportunities for biomarker discovery and disease monitoring in AD.

## Data Availability

All data are available upon request

## Author Contributions

DC designed the study, performed experiments, analysed data, and wrote the manuscript. IVB and GB analysed data and wrote the manuscript, SRP and PLO performed experiments, TPV analysed data, LP enrolled patients, and provided CSF samples, CRJ designed the study and contributed in writing the manuscript. All authors revised and edited the manuscript.

## Acknowledgments

VUmc-Cancer Center Amsterdam is acknowledged for the proteomics infrastructure. The authors wish to thank Dr. Federica Susta and Cristiano Spaccatini for assisting in handling of CSF samples.

## References

(1) Raposo, G.; Stoorvogel, W. Extracellular Vesicles: Exosomes, Microvesicles, and Friends. Journal of Cell Biology. 2013. https://doi.org/10.1083/jcb.201211138.

(2) Tkach, M.; Théry, C. Communication by Extracellular Vesicles: Where We Are and Where We Need to Go. Cell. 2016. https://doi.org/10.1016/j.cell.2016.01.043.

(3) Colombo, M.; Raposo, G.; Théry, C. Biogenesis, Secretion, and Intercellular Interactions of Exosomes and Other Extracellular Vesicles. Annu. Rev. Cell Dev. Biol. 2014, 30 (1), 255–289. https://doi.org/10.1146/annurev-cellbio-101512-122326.

(4) Sharma, P.; Schiapparelli, L.; Cline, H. T. Exosomes Function in Cell-Cell Communication during Brain Circuit Development. Current Opinion in Neurobiology. 2013. https://doi.org/10.1016/j.conb.2013.08.005.

(5) Janas, A. M.; Sapoń, K.; Janas, T.; Stowell, M. H. B.; Janas, T. Exosomes and Other Extracellular Vesicles in Neural Cells and Neurodegenerative Diseases. Biochimica et Biophysica Acta - Biomembranes. 2016. https://doi.org/10.1016/j.bbamem.2016.02.011.

(6) Lim, Y. J.; Lee, S. J. Are Exosomes the Vehicle for Protein Aggregate Propagation in Neurodegenerative Diseases? Acta neuropathologica communications. 2017. https://doi.org/10.1186/s40478-017-0467-z.

(7) Howitt, J.; Hill, A. F. Exosomes in the Pathology of Neurodegenerative Diseases. Journal of Biological Chemistry. 2016. https://doi.org/10.1074/jbc.R116.757955.

(8) Molinuevo, J. L.; Ayton, S.; Batrla, R.; Bednar, M. M.; Bittner, T.; Cummings, J.; Fagan, A. M.; Hampel, H.; Mielke, M. M.; Mikulskis, A.; et al. Current State of Alzheimer’s Fluid Biomarkers. Acta Neuropathol. 2018. https://doi.org/10.1007/s00401-018-1932-x.

(9) Parnetti, L.; Gaetani, L.; Eusebi, P.; Paciotti, S.; Hansson, O.; El-Agnaf, O.; Mollenhauer, B.; Blennow, K.; Calabresi, P. CSF and Blood Biomarkers for Parkinson’s Disease. The Lancet Neurology. 2019. https://doi.org/10.1016/S1474-4422(19)30024-9.

(10) Emelyanov, A.; Shtam, T.; Kamyshinsky, R.; Garaeva, L.; Verlov, N.; Miliukhina, I.; Kudrevatykh, A.; Gavrilov, G.; Zabrodskaya, Y.; Pchelina, S.; et al. Cryo-Electron Microscopy of Extracellular Vesicles from Cerebrospinal Fluid. PLoS One 2020, 15 (1), e0227949. https://doi.org/10.1371/journal.pone.0227949.

(11) Chiasserini, D.; Van Weering, J. R. T.; Piersma, S. R.; Pham, T. V.; Malekzadeh, A.; Teunissen, C. E.; De Wit, H.; Jiménez, C. R. Proteomic Analysis of Cerebrospinal Fluid Extracellular Vesicles: A Comprehensive Dataset. J. Proteomics 2014, 106, 191–204. https://doi.org/10.1016/j.jprot.2014.04.028.

(12) Street, J. M.; Barran, P. E.; Mackay, C. L.; Weidt, S.; Balmforth, C.; Walsh, T. S.; Chalmers, R. T. A.; Webb, D. J.; Dear, J. W. Identification and Proteomic Profiling of Exosomes in Human Cerebrospinal Fluid. J. Transl. Med. 2012. https://doi.org/10.1186/1479-5876-10-5.

(13) Théry, C.; Witwer, K. W.; Aikawa, E.; Alcaraz, M. J.; Anderson, J. D.; Andriantsitohaina, R.; Antoniou, A.; Arab, T.; Archer, F.; Atkin-Smith, G. K.; et al. Minimal Information for Studies of Extracellular Vesicles 2018 (MISEV2018): A Position Statement of the International Society for Extracellular Vesicles and Update of the MISEV2014 Guidelines. J. Extracell. Vesicles 2018. https://doi.org/10.1080/20013078.2018.1535750.

(14) Kowal, J.; Arras, G.; Colombo, M.; Jouve, M.; Morath, J. P.; Primdal-Bengtson, B.; Dingli, F.; Loew, D.; Tkach, M.; Théry, C. Proteomic Comparison Defines Novel Markers to Characterize Heterogeneous Populations of Extracellular Vesicle Subtypes. Proc. Natl. Acad. Sci. U. S. A. 2016. https://doi.org/10.1073/pnas.1521230113.

(15) Lee, S.; Mankhong, S.; Kang, J. H. Extracellular Vesicle as a Source of Alzheimer’s Biomarkers: Opportunities and Challenges. International Journal of Molecular Sciences. 2019. https://doi.org/10.3390/ijms20071728.

(16) Sardar Sinha, M.; Ansell-Schultz, A.; Civitelli, L.; Hildesjö, C.; Larsson, M.; Lannfelt, L.; Ingelsson, M.; Hallbeck, M. Alzheimer’s Disease Pathology Propagation by Exosomes Containing Toxic Amyloid-Beta Oligomers. Acta Neuropathol. 2018. https://doi.org/10.1007/s00401-018-1868-1.

(17) Asai, H.; Ikezu, S.; Tsunoda, S.; Medalla, M.; Luebke, J.; Haydar, T.; Wolozin, B.; Butovsky, O.; Kügler, S.; Ikezu, T. Depletion of Microglia and Inhibition of Exosome Synthesis Halt Tau Propagation. Nat. Neurosci. 2015. https://doi.org/10.1038/nn.4132.

(18) Spitzer, P.; Mulzer, L. M.; Oberstein, T. J.; Munoz, L. E.; Lewczuk, P.; Kornhuber, J.; Herrmann, M.; Maler, J. M. Microvesicles from Cerebrospinal Fluid of Patients with Alzheimer’s Disease Display Reduced Concentrations of Tau and APP Protein. Sci. Rep. 2019. https://doi.org/10.1038/s41598-019-43607-7.

(19) Pulliam, L.; Sun, B.; Mustapic, M.; Chawla, S.; Kapogiannis, D. Plasma Neuronal Exosomes Serve as Biomarkers of Cognitive Impairment in HIV Infection and Alzheimer’s Disease. J. Neurovirol. 2019. https://doi.org/10.1007/s13365-018-0695-4.

(20) Saman, S.; Kim, W. H.; Raya, M.; Visnick, Y.; Miro, S.; Saman, S.; Jackson, B.; McKee, A. C.; Alvarez, V. E.; Lee, N. C. Y.; et al. Exosome-Associated Tau Is Secreted in Tauopathy Models and Is Selectively Phosphorylated in Cerebrospinal Fluid in Early Alzheimer Disease. J. Biol. Chem. 2012. https://doi.org/10.1074/jbc.M111.277061.

(21) Piersma, S. R.; Warmoes, M. O.; de Wit, M.; de Reus, I.; Knol, J. C.; Jiménez, C. R. Whole Gel Processing Procedure for GeLC-MS/MS Based Proteomics. Proteome Sci. 2013, 11, 17. https://doi.org/10.1186/1477-5956-11-17.

(22) Cox, J.; Mann, M. MaxQuant Enables High Peptide Identification Rates, Individualized p.p.b.-Range Mass Accuracies and Proteome-Wide Protein Quantification. Nat. Biotechnol. 2008, 26, 1367–1372. https://doi.org/10.1038/nbt.1511.

(23) R Core Team 2019. R: A Language and Environment for Statistical Computing. R Foundation for Statistical Computing, Vienna, Austria. URL http://www.R-project.org/. 2019.

(24) Schwanhüusser, B.; Busse, D.; Li, N.; Dittmar, G.; Schuchhardt, J.; Wolf, J.; Chen, W.; Selbach, M. Global Quantification of Mammalian Gene Expression Control. Nature 2011. https://doi.org/10.1038/nature10098.

(25) Ritchie, M. E.; Phipson, B.; Wu, D.; Hu, Y.; Law, C. W.; Shi, W.; Smyth, G. K. Limma Powers Differential Expression Analyses for RNA-Sequencing and Microarray Studies. Nucleic Acids Res. 2015. https://doi.org/10.1093/nar/gkv007.

(26) Wei, R.; Wang, J.; Su, M.; Jia, E.; Chen, S.; Chen, T.; Ni, Y. Missing Value Imputation Approach for Mass Spectrometry-Based Metabolomics Data. Sci. Rep. 2018. https://doi.org/10.1038/s41598-017-19120-0.

(27) Yu, G.; Wang, L. G.; Han, Y.; He, Q. Y. ClusterProfiler: An R Package for Comparing Biological Themes among Gene Clusters. Omi. A J. Integr. Biol. 2012. https://doi.org/10.1089/omi.2011.0118.

(28) Subramanian, A.; Tamayo, P.; Mootha, V. K.; Mukherjee, S.; Ebert, B. L.; Gillette, M. A.; Paulovich, A.; Pomeroy, S. L.; Golub, T. R.; Lander, E. S.; et al. Gene Set Enrichment Analysis: A Knowledge-Based Approach for Interpreting Genome-Wide Expression Profiles. Proc. Natl. Acad. Sci. U. S. A. 2005. https://doi.org/10.1073/pnas.0506580102.

(29) Liberzon, A.; Birger, C.; Thorvaldsdóttir, H.; Ghandi, M.; Mesirov, J. P.; Tamayo, P. The Molecular Signatures Database Hallmark Gene Set Collection. Cell Syst. 2015. https://doi.org/10.1016/j.cels.2015.12.004.

(30) Szklarczyk, D.; Gable, A. L.; Lyon, D.; Junge, A.; Wyder, S.; Huerta-Cepas, J.; Simonovic, M.; Doncheva, N. T.; Morris, J. H.; Bork, P.; et al. STRING V11: Protein-Protein Association Networks with Increased Coverage, Supporting Functional Discovery in Genome-Wide Experimental Datasets. Nucleic Acids Res. 2019. https://doi.org/10.1093/nar/gky1131.

(31) Shannon, P.; Markiel, A.; Ozier, O.; Baliga, N. S.; Wang, J. T.; Ramage, D.; Amin, N.; Schwikowski, B.; Ideker, T. Cytoscape□: A Software Environment for Integrated Models of Biomolecular Interaction Networks Cytoscape□: A Software Environment for Integrated Models of Biomolecular Interaction Networks. Genome Res. 2003. https://doi.org/10.1101/gr.1239303.

(32) Enright, A. J. An Efficient Algorithm for Large-Scale Detection of Protein Families. Nucleic Acids Res. 2002. https://doi.org/10.1093/nar/30.7.1575.

(33) Morris, J. H.; Apeltsin, L.; Newman, A. M.; Baumbach, J.; Wittkop, T.; Su, G.; Bader, G. D.; Ferrin, T. E. ClusterMaker: A Multi-Algorithm Clustering Plugin for Cytoscape. BMC Bioinformatics 2011, 12 (1), 436. https://doi.org/10.1186/1471-2105-12-436.

(34) Buzás, E. I.; Tóth, E.; Sódar, B. W.; Szabó-Taylor, K. Molecular Interactions at the Surface of Extracellular Vesicles. Seminars in Immunopathology. 2018. https://doi.org/10.1007/s00281-018-0682-0.

(35) Uhlén, M.; Fagerberg, L.; Hallström, B. M.; Lindskog, C.; Oksvold, P.; Mardinoglu, A.; Sivertsson, Å., Kampf, C.; Sjöstedt, E.; Asplund, A.; et al. Tissue-Based Map of the Human Proteome. Science (80-.). 2015. https://doi.org/10.1126/science.1260419.

(36) Perrotte, M.; Haddad, M.; Le Page, A.; Frost, E. H.; Fulöp, T.; Ramassamy, C. Profile of Pathogenic Proteins in Total Circulating Extracellular Vesicles in Mild Cognitive Impairment and during the Progression of Alzheimer’s Disease. Neurobiol. Aging 2019. https://doi.org/10.1016/j.neurobiolaging.2019.10.010.

(37) Guix, F. X.; Corbett, G. T.; Cha, D. J.; Mustapic, M.; Liu, W.; Mengel, D.; Chen, Z.; Aikawa, E.; Young-Pearse, T.; Kapogiannis, D.; et al. Detection of Aggregation-Competent Tau in Neuron-Derived Extracellular Vesicles. Int. J. Mol. Sci. 2018. https://doi.org/10.3390/ijms19030663.

(38) Stuendl, A.; Kunadt, M.; Kruse, N.; Bartels, C.; Moebius, W.; Danzer, K. M.; Mollenhauer, B.; Schneider, A. Induction of α-Synuclein Aggregate Formation by CSF Exosomes from Patients with Parkinson’s Disease and Dementia with Lewy Bodies. Brain 2016. https://doi.org/10.1093/brain/awv346.

(39) Hondius, D. C.; van Nierop, P.; Li, K. W.; Hoozemans, J. J. M.; van der Schors, R. C.; van Haastert, E. S.; van der Vies, S. M.; Rozemuller, A. J. M.; Smit, A. B. Profiling the Human Hippocampal Proteome at All Pathologic Stages of Alzheimer’s Disease. Alzheimers. Dement. 2016. https://doi.org/10.1016/j.jalz.2015.11.002.

(40) Bai, B.; Wang, X.; Li, Y.; Chen, P.-C.; Yu, K.; Dey, K. K.; Yarbro, J. M.; Han, X.; Lutz, B. M.; Rao, S.; et al. Deep Multilayer Brain Proteomics Identifies Molecular Networks in Alzheimer’s Disease Progression. Neuron 2020. https://doi.org/10.1016/j.neuron.2019.12.015.

(41) Olsson, B.; Lautner, R.; Andreasson, U.; Öhrfelt, A.; Portelius, E.; Bjerke, M.; Hölttä, M.; Rosen, C.; Dakin, K.; Wu, E.; et al. Alzbiomarker Database on Established and Novel CSF and Plasma Biomarkers for Alzheimer’s Disease. Alzheimer’s Dement. 2015. https://doi.org/10.1016/j.jalz.2015.07.422.

(42) Jahanshad, N.; Rajagopalan, P.; Hua, X.; Hibar, D. P.; Nir, T. M.; Toga, A. W.; Jack, C. R.; Saykin, A. J.; Green, R. C.; Weiner, M. W.; et al. Genome-Wide Scan of Healthy Human Connectome Discovers SPON1 Gene Variant Influencing Dementia Severity. Proc. Natl. Acad. Sci. U. S. A. 2013. https://doi.org/10.1073/pnas.1216206110.

(43) Parnetti, L.; Chiasserini, D.; Eusebi, P.; Giannandrea, D.; Bellomo, G.; De Carlo, C.; Padiglioni, C.; Mastrocola, S.; Lisetti, V.; Calabresi, P. Performance of Aβ1-40, Aβ1-42, Total Tau, and Phosphorylated Tau as Predictors of Dementia in a Cohort of Patients with Mild Cognitive Impairment. J. Alzheimer’s Dis. 2012, 29, 229–238. https://doi.org/10.3233/JAD-2011-111349.

(44) McCullough, J.; Colf, L. A.; Sundquist, W. I. Membrane Fission Reactions of the Mammalian ESCRT Pathway. Annu. Rev. Biochem. 2013. https://doi.org/10.1146/annurev-biochem-072909-101058.

(45) Baietti, M. F.; Zhang, Z.; Mortier, E.; Melchior, A.; Degeest, G.; Geeraerts, A.; Ivarsson, Y.; Depoortere, F.; Coomans, C.; Vermeiren, E.; et al. Syndecan-Syntenin-ALIX Regulates the Biogenesis of Exosomes. Nat. Cell Biol. 2012. https://doi.org/10.1038/ncb2502.

(46) Brown, P. D.; Davies, S. L.; Speake, T.; Millar, I. D. Molecular Mechanisms of Cerebrospinal Fluid Production. Neuroscience. 2004, pp 957–970. https://doi.org/10.1016/j.neuroscience.2004.07.003.

(47) Damkier, H. H.; Brown, P. D.; Praetorius, J. Cerebrospinal Fluid Secretion by the Choroid Plexus. Physiological Reviews. 2013. https://doi.org/10.1152/physrev.00004.2013.

(48) Gorivodsky, M.; Mukhopadhyay, M.; Wilsch-Braeuninger, M.; Phillips, M.; Teufel, A.; Kim, C.; Malik, N.; Huttner, W.; Westphal, H. Intraflagellar Transport Protein 172 Is Essential for Primary Cilia Formation and Plays a Vital Role in Patterning the Mammalian Brain. Dev. Biol. 2009. https://doi.org/10.1016/j.ydbio.2008.09.019.

(49) Cantagrel, V.; Silhavy, J. L.; Bielas, S. L.; Swistun, D.; Marsh, S. E.; Bertrand, J. Y.; Audollent, S.; Attié-Bitach, T.; Holden, K. R.; Dobyns, W. B.; et al. Mutations in the Cilia Gene ARL13B Lead to the Classical Form of Joubert Syndrome. Am. J. Hum. Genet. 2008. https://doi.org/10.1016/j.ajhg.2008.06.023.

(50) Alkanderi, S.; Molinari, E.; Shaheen, R.; Elmaghloob, Y.; Stephen, L. A.; Sammut, V.; Ramsbottom, S. A.; Srivastava, S.; Cairns, G.; Edwards, N.; et al. ARL3 Mutations Cause Joubert Syndrome by Disrupting Ciliary Protein Composition. Am. J. Hum. Genet. 2018. https://doi.org/10.1016/j.ajhg.2018.08.015.

(51) Balusu, S.; Van Wonterghem, E.; De Rycke, R.; Raemdonck, K.; Stremersch, S.; Gevaert, K.; Brkic, M.; Demeestere, D.; Vanhooren, V.; Hendrix, A.; et al. Identification of a Novel Mechanism of Blood–Brain Communication during Peripheral Inflammation via Choroid Plexus□derived Extracellular Vesicles. EMBO Mol. Med. 2016. https://doi.org/10.15252/emmm.201606271.

(52) Gul, I. S.; Hulpiau, P.; Saeys, Y.; van Roy, F. Evolution and Diversity of Cadherins and Catenins. Experimental Cell Research. 2017. https://doi.org/10.1016/j.yexcr.2017.03.001.

(53) Tietz, S.; Engelhardt, B. Brain Barriers: Crosstalk between Complex Tight Junctions and Adherens Junctions. Journal of Cell Biology. 2015. https://doi.org/10.1083/jcb.201412147.

(54) Li, W.; Chen, Z.; Chin, I.; Chen, Z.; Dai, H. The Role of VE-Cadherin in Blood-Brain Barrier Integrity Under Central Nervous System Pathological Conditions. Curr. Neuropharmacol. 2018. https://doi.org/10.2174/1570159x16666180222164809.

(55) Herradõn, G.; Pérez-García, C. Targeting Midkine and Pleiotrophin Signalling Pathways in Addiction and Neurodegenerative Disorders: Recent Progress and Perspectives. British Journal of Pharmacology. 2014. https://doi.org/10.1111/bph.12312.

(56) Michibata, H.; Okuno, T.; Konishi, N.; Kyono, K.; Wakimoto, K.; Aoki, K.; Kondo, Y.; Takata, K.; Kitamura, Y.; Taniguchi, T. Human GPM6A Is Associated with Differentiation and Neuronal Migration of Neurons Derived from Human Embryonic Stem Cells. Stem Cells Dev. 2009, 18 (4), 629–639. https://doi.org/10.1089/scd.2008.0215.

(57) Muraoka, S.; DeLeo, A. M.; Sethi, M. K.; Yukawa-Takamatsu, K.; Yang, Z.; Ko, J.; Hogan, J. D.; Daley, S. A.; Ruan, Z.; You, Y.; et al. Proteomic Profiling and Biological Characterization of Extracellular Vesicles Isolated from Alzheimer’s Disease Brain Tissues. bioRxiv 2019. https://doi.org/10.1101/733477.

(58) Sogorb-Esteve, A.; García-Ayllón, M. S.; Gobom, J.; Alom, J.; Zetterberg, H.; Blennow, K.; Sáez-Valero, J. Levels of ADAM10 Are Reduced in Alzheimer’s Disease CSF. J. Neuroinflammation 2018. https://doi.org/10.1186/s12974-018-1255-9.

(59) Craig-Schapiro, R.; Perrin, R. J.; Roe, C. M.; Xiong, C.; Carter, D.; Cairns, N. J.; Mintun, M. A.; Peskind, E. R.; Li, G.; Galasko, D. R.; et al. YKL-40: A Novel Prognostic Fluid Biomarker for Preclinical Alzheimer’s Disease. Biol. Psychiatry 2010. https://doi.org/10.1016/j.biopsych.2010.08.025.

(60) Falcon, C.; Monté-Rubio, G. C.; Grau-Rivera, O.; Suárez-Calvet, M.; Sánchez-Valle, R.; Rami, L.; Bosch, B.; Haass, C.; Gispert, J. D.; Molinuevo, J. L. CSF Glial Biomarkers YKL40 and STREM2 Are Associated with Longitudinal Volume and Diffusivity Changes in Cognitively Unimpaired Individuals. NeuroImage Clin. 2019. https://doi.org/10.1016/j.nicl.2019.101801.

(61) Bonneh-Barkay, D.; Wang, G.; Starkey, A.; Hamilton, R. L.; Wiley, C. A. In Vivo CHI3L1 (YKL-40) Expression in Astrocytes in Acute and Chronic Neurological Diseases. J. Neuroinflammation 2010. https://doi.org/10.1186/1742-2094-7-34.

(62) del Campo, M.; Galimberti, D.; Elias, N.; Boonkamp, L.; Pijnenburg, Y. A.; van Swieten, J. C.; Watts, K.; Paciotti, S.; Beccari, T.; Hu, W.; et al. Novel CSF Biomarkers to Discriminate FTLD and Its Pathological Subtypes. Ann. Clin. Transl. Neurol. 2018. https://doi.org/10.1002/acn3.629.

(63) Klar, A.; Baldassare, M.; Jessell, T. M. F-Spondin: A Gene Expressed at High Levels in the Floor Plate Encodes a Secreted Protein That Promotes Neural Cell Adhesion and Neurite Extension. Cell 1992. https://doi.org/10.1016/0092-8674(92)90121-R.

(64) Xiong, F.; Ge, W.; Ma, C. Quantitative Proteomics Reveals Distinct Composition of Amyloid Plaques in Alzheimer’s Disease. Alzheimer’s Dement. 2019. https://doi.org/10.1016/j.jalz.2018.10.006.

(65) Yuyama, K.; Igarashi, Y. Exosomes as Carriers of Alzheimer’s Amyloid-SS. Front. Neurosci. 2017. https://doi.org/10.3389/fnins.2017.00229.

(66) Allison, D. B.; Cui, X.; Page, G. P.; Sabripour, M. Microarray Data Analysis: From Disarray to Consolidation and Consensus. Nature Reviews Genetics. 2006. https://doi.org/10.1038/nrg1749.

(67) Bai, B.; Hales, C. M.; Chen, P. C.; Gozal, Y.; Dammer, E. B.; Fritz, J. J.; Wang, X.; Xia, Q.; Duong, D. M.; Street, C.; et al. U1 Small Nuclear Ribonucleoprotein Complex and RNA Splicing Alterations in Alzheimer’s Disease. Proc. Natl. Acad. Sci. U. S. A. 2013. https://doi.org/10.1073/pnas.1310249110.

(68) Zhang, P.; Zhou, X.; He, M.; Shang, Y.; Tetlow, A. L.; Godwin, A. K.; Zeng, Y. Ultrasensitive Detection of Circulating Exosomes with a 3D-Nanopatterned Microfluidic Chip. Nat. Biomed. Eng. 2019. https://doi.org/10.1038/s41551-019-0356-9.

(69) Winston, C. N.; Romero, H. K.; Ellisman, M.; Nauss, S.; Julovich, D. A.; Conger, T.; Hall, J. R.; Campana, W.; O’Bryant, S. E.; Nievergelt, C. M.; et al. Assessing Neuronal and Astrocyte Derived Exosomes From Individuals With Mild Traumatic Brain Injury for Markers of Neurodegeneration and Cytotoxic Activity. Front. Neurosci. 2019. https://doi.org/10.3389/fnins.2019.01005.

(70) Bijnsdorp, I. V.; Maxouri, O.; Kardar, A.; Schelfhorst, T.; Piersma, S. R.; Pham, T. V.; Vis, A.; Van Moorselaar, R. J.; Jimenez, C. R. Feasibility of Urinary Extracellular Vesicle Proteome Profiling Using a Robust and Simple, Clinically Applicable Isolation Method. J. Extracell. Vesicles 2017, 6 (1). https://doi.org/10.1080/20013078.2017.1313091.

